# Detection and quantification of human immunodeficiency virus-1 (HIV-1) total nucleic acids in wastewater settled solids from two California communities

**DOI:** 10.1101/2024.03.12.24304178

**Authors:** Marlene K. Wolfe, Bridgette Shelden, Dorothea Duong, Meri R.J. Varkila, Bradley J. White, Julie Parsonnet, Alexandria B. Boehm

## Abstract

Wastewater surveillance for infectious agents has proved useful in identifying circulation of viruses within populations. We investigated the presence and concentration of human immunodeficiency virus (HIV)-1 total nucleic acids (including both viral RNA and proviral DNA) in wastewater solids. We retrospectively measured HIV-1 nucleic-acids in two samples per week for 26 months at two wastewater treatment plants serving populations with different prevalences of HIV infections in San Francisco and Santa Clara County, California, USA. We detected HIV nucleic-acids in a majority of samples with concentrations ranging from non-detect to 3.9×10^5^ cp/g (N=459 samples total). Concentrations of HIV-1 were significantly higher in samples from the wastewater treatment plant serving a population with a higher prevalence of people living with HIV than in the plant serving a population with lower prevalence. The HIV-1 nucleic-acids amplified were primarily DNA and thus represented proviral DNA shedding into wastewater. Additionally, we found that HIV-1 nucleic-acid concentrations in wastewater solids were orders of magnitude higher than those in liquid wastewater indicating that the HIV-1 target preferentially sorbs to solids. Whether concentrations of HIV-1 in wastewater solids can be used to identify numbers of incident cases remains unknown. Additional work on HIV-1 shedding from individuals with viremia and people living with HIV is needed to translate wastewater measurements to quantitative information on infections.

## Introduction

Despite advances in prevention and treatment and a decrease in new human immunodeficiency virus (HIV) infections, more than 600,000 people die from acquired immunodeficiency syndrome (AIDS) annually^1^. The Ending the HIV Epidemic in the United States (EHE) initiative has set an ambitious goal of reducing the number of new HIV infections in the US by 90% between 2019 and 2030. Progress, however, has been below target and disproportionately slow in some demographic groups and geographic regions. At the end of 2021, there were an estimated 1.2 million people living with HIV (PLWH) in the US and 36,136 people who were newly diagnosed^2,3^ [ref].

Minoritized people are disproportionately affected by the HIV epidemic in the US; 40% of people receiving a new diagnosis in the US in 2021 were Black^2^. Although it has been estimated that 87% of those living with HIV in the US are aware of their status, during the COVID-19 pandemic, HIV testing and new diagnoses decreased. Consequently, a greater proportion of PLWH may be unaware of their status and the CDC has identified specific geographic regions where underdiagnosis may be particularly problematic ^4–7^. Systemic barriers to care, exacerbated by the pandemic, may continue to slow progress towards EHE goals. Ongoing assessment of the prevalence of HIV infection in communities, especially those underserved by healthcare and testing centers, is critical to provide resources that ensure consistent testing and subsequent treatment.

Wastewater monitoring has not been previously applied to HIV but could help identify trends in infection prevalence and identify pockets of underdiagnosed infection. Wastewater, a naturally composited biological sample that includes bodily secretions, has been used to monitor a wide range of infectious diseases including respiratory viruses, enteric viruses, dengue (an arbovirus) and mpox (a poxvirus).^8–14^ Wastewater monitoring was effectively used to support mpox response at the start of the outbreak in the United States during summer 2022^10^; providing information to respond to an outbreak that was syndemic with HIV.

Although the use of wastewater monitoring for HIV has not been explored, evidence from early in the HIV pandemic suggests that viral RNA and DNA can be detected in wastewater. Several studies from the early 1990’s demonstrated the detection of HIV nucleic acids and intact virus in natural wastewaters^15,16^ and detectability of the virus seeded in wastewater^17,18^. These studies were focused on occupational hazard, rather than monitoring. Some data indicate that HIV nucleic acids are shed in urine and feces of people living with HIV (PLWH), even those who are virally suppressed ^19–21^. Contributions to wastewater therefore may be expected from people living with HIV. This suggests that detection and quantification for monitoring of HIV infections at the community level is a realistic possibility.

The goal of this study is to apply testing for HIV RNA and DNA to wastewater samples from two adjacent counties to determine whether consistent detection and quantification of HIV nucleic acids in wastewater is feasible, and explore the relationship of these measurements to the overall estimated rates of people living with HIV in the associated communities.

## Methods

### Study design and sample collection

We conducted a longitudinal retrospective analysis of HIV nucleic acids in wastewater solids from two wastewater treatment plants (WWTPs) in the San Francisco Bay Area, California (CA), USA. Fifty milliliters (50 ml) samples of wastewater solids were collected from primary clarifiers daily using sterile methods, and biobanked with consent of the participating WWTPs as part of a routine wastewater monitoring program that began in November 2020. Two samples per week covering a period of 26 months were selected for this study. They were collected between 2 Feb 2021 and 14 April 2023 (459 samples total). The two WWTPs included in the retrospective analysis serve ∼75% (1,500,000 people) of Santa Clara County, (SJ) and ∼25% (250,000 people) of San Francisco County (OSP), CA. Further WWTP descriptions are elsewhere^22^. These sites were selected and tested as part of a retrospective study measuring nucleic acids from a panel of viruses that are candidates for wastewater monitoring; HIV-1 was one of the targets selected for inclusion in this study. Samples were stored at 4°C, transported to the lab, and processed within six hours. Solids were then dewatered using centrifugation^23^ and then frozen at -80°C for 4 - 60 weeks.

We collected additional samples over a consecutive 9 days to compare concentrations of HIV-1 between liquid wastewater and wastewater solids, and to compare measurements made with and without a reverse-transcription step during the analytic analysis. Additional samples were collected from two sites, OSP and a second site in San Francisco County (SEP) that serves ∼75% of the county (650,000 people). For these two sites, 24-hour composite samples of raw wastewater influent (liquid) and a grab sample of solids from the primary clarifiers were collected on the same day for 8 days in early July 2023, and one additional solids samples was collected from SEP (Table S1, samples hereafter labeled 1-8 for OSP and SEP liquid, and 1-8 and 1-9 for OSP and SEP solids, respectively). All the solids samples (n=17) were tested for HIV-1 target using both RT-PCR and PCR to gain insight into the proportion of HIV-1 RNA and DNA in the samples, respectively (Table S1). Samples were stored at 4°C for the duration of the 9 day study and then underwent pre-analytical and analytical processing without any additional storage. Solids were dewatered immediately using centrifugation^23^ and subsequently processed for nucleic-acid extraction without storage.

### Solids pre-analytical methods

Frozen, dewatered solids were thawed overnight at 4°C. Solids were resuspended in DNA/RNA Shield (Zymo Research, Irvine, CA) at a concentration of approximately 0.75 mg (wet weight)/ml. This concentration was chosen as it was shown to reduce inhibition in RT-PCR applications with extracted nucleic acids^24^. A separate aliquot of dewatered solids was dried in an oven to determine its dry weight, and the calculated percent solids was used to express concentrations of nucleic acid targets in units of per gram dry weight. These pre-analytical methods are described thoroughly in a publicly accessible protocol^25^. Nucleic-acids were extracted from 10 replicate aliquots of dewatered settled solids suspended in the DNA/RNA Shield using the Chemagic Viral DNA/RNA 300 kit H96 for the Perkin Elmer Chemagic 360 (Perkin Elmer, Waltham, MA) followed by PCR inhibitor removal with the Zymo OneStep-96 PCR Inhibitor Removal kit (Zymo Research, Irvine, CA). 300 μl of the suspension entered into the nucleic-acid extraction process and 50 μl of nucleic-acids are retrieved after the inhibitor removal kit. Negative extraction controls consisted of BCoV-vaccine spiked DNA/RNA Shield. Nucleic acid extracts for the biobanked wastewater samples used in the retrospective study were stored at -80°C for different amounts of time (8-273 days, median = 266 days) and subjected to one freeze thaw prior to analysis. Nucleic-acids from the solids samples used for the liquid/solid comparison and with/without reverse-transcription were not stored prior to analysis.

### Liquids pre-analytical methods

Viral particles were concentrated from wastewater using an affinity-based capture method with magnetic hydrogel Nanotrap Particles (Ceres Nanosciences, Manassas, VA). Samples were spiked with BCoV before concentration, and used to assess nucleic acid recovery. For each sample, 10 replicate aliquots of 10 mL of liquid wastewater were concentrated using the Nanotrap Particles and Enhancement Reagent 1 on a KingFisher Flex system. Nucleic acids were then extracted from each replicate with the MagMAX Viral/Pathogen Nucleic Acid Isolation Kit (Applied Biosystems, Waltham, MA) using the KingFisher Flex system. Resulting nucleic acids were then processed through a Zymo OneStep-96 PCR Inhibitor Removal kit (Zymo Research, Irvine, CA). Each 10 ml sample resulted in 50 μl total nucleic acid extract. Note that suspended solids were not removed prior to initiating these methods, and the nucleic-acids were not stored prior to analysis.

### HIV (Reverse-transcription)-polymerase chain reaction assay choice

We selected previously published HIV genome-specific primers and probes targeting the long terminal repeat (LTR) region of the HIV-1 genome for use in this study; the assay chosen does not target HIV-2^26^. We used the 496F/546P/622R primers and probe as recommended by Kibirige et al.^27^ The assay uses primers and probes originally described in Brussels et al.^28^ and Friedrich et al. ^29^ (Table 1).

**Table 1.**
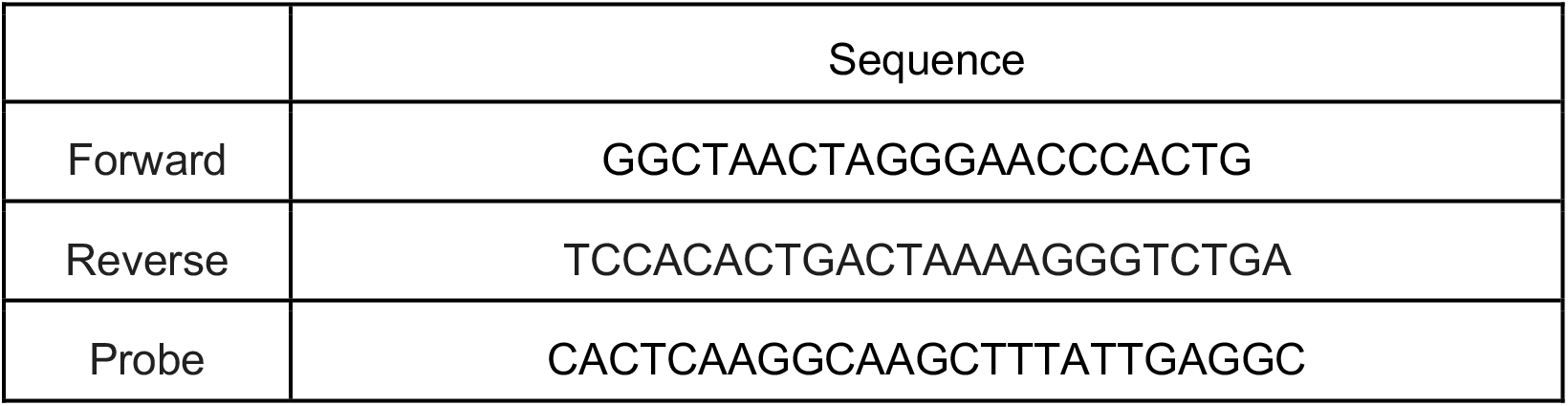
Primer and hydrolysis probes targeting the long terminal report (LTR) region of HIV-1. The amplicon size is 127 basepairs (bp). The HIV-1 probe contained Cyanine-5 (Cy5) fluorescent molecule, ZEN, a proprietary internal quencher from Integrated DNA Technologies (Coralville, IA, USA); and IBFQ, Iowa Black FQ.

To confirm HIV-1 assay specificity, the assay was challenged with non-target viruses including those in two panels purchased from Zeptomatrix (Buffalo, NY), NATRVP2.1-BIO and NATEVP-C, along with HIV-2 (ATCC PTA-9773). The NATRVP2.1-BIO panel includes chemically inactivated intact influenza viruses, parainfluenza viruses, adenovirus, rhinovirus, metapneumovirus, and coronaviruses. The NATEVP-C panel includes chemically inactivated intact coxsackieviruses, echovirus, and parechovirus. To confirm HIV-1 assay sensitivity, synthetic HIV-1 RNA (ATCC VR-3351SD) was used as a positive control for HIV-1. American Type Culture Collection (ATCC) controls are synthetic RNA. Nucleic-acids (NA) were extracted from intact viruses using the Chemagic Viral DNA/RNA 300 kit H96 (PerkinElmer, Waltham, MA).

### Digital-droplet RT-PCR methods

For sensitivity and specificity testing of the HIV-1 assay, nucleic-acids were used undiluted as template in digital RT-PCR reactions in a single well. No template negative RT-PCR controls using molecular grade water as template were included in each plate. The assay was run in simplex with only primers and probe in Table 1.

Wastewater samples were tested for HIV-1 nucleic-acids as well as pepper mild mottle virus (PMMoV) using droplet digital RT-PCR. PMMoV is a pepper virus that is highly abundant in wastewater across the world and used in this study as an endogenous recovery control, as well as a measure of fecal strength of the wastewater^30^. We did not utilize an exogenous extraction control (such as a spiked virus like bovine coronavirus) due to the difficulty in interpreting recovery with an exogenous target^31^; however this method has consistently provided high recoveries approaching 100% of spiked BCoV in our laboratories^22^.

Each of the 10 replicate nucleic-acid wastewater extracts was used as template undiluted in its own well (10 wells per sample) to measure HIV-1 and PMMoV. The HIV-1 assay was run in multiplex using a probe mixing approach with seven other assays targeting genomes of rotavirus, influenza A subtype markers H1 and N1, SARS-CoV-2, human norovirus GII, human adenovirus group F, and West Nile virus (results not reported herein, but provided elsewhere^32,33^). Multiplexing method testing showed that presence of seven other nucleic-acid targets in the multiplex reaction did not affect quantification of the HIV-1 target (Figure S1).

Nucleic-acids were diluted 1:100 and used as template to measure PMMoV using methods outlined in detail elsewhere^22^ and thoroughly in a protocols.io protocol^34^ (Table S2 provides primers and probe). Extraction negative (DNA/RNA shield, 3 wells) controls, and PCR negative (molecular grade water, 3 wells) and positive controls (synthetic RNA, 1 well) were run on each 96 well plate. Any failure of these controls resulted in re-processing of the samples, and any samples with anomalously low PMMoV would also be rerun or discarded from analysis.

ddRT-PCR was performed on 20 μl samples from a 22 μl reaction volume, prepared using 5.5 μl template, mixed with 5.5 μl of One-Step RT-ddPCR Advanced Kit for Probes (Bio-Rad 1863021), 2.2 μl of 200 U/μl Reverse Transcriptase, 1.1 μl of 300 mM dithiothreitol (DTT) and primers and probes mixtures at a final concentration of 900 nM and 250 nM respectively. Primer and probes for assays were purchased from Integrated DNA Technologies (IDT, San Diego, CA) (Table 1 and S2).

HIV-1 nucleic-acids were quantified in the 17 solids samples collected at OSP and SEP in July 2023 (Table S1) using both RT-PCR and PCR without the RT step. RT-PCR was run as described above for the biobanked wastewater samples. PCR was run using the same method as for RT-PCR but the RT enzyme and DTT were omitted from the mastermix. As a control, the concentration of the N gene of SARS-CoV-2 was examined from the multiplex assay as the N gene is expected to only be detected when an RT step is included as it resides in wastewater as genomic RNA. We confirmed that the N gene was not detected without the RT step indicating lack of RT activity in that mastermix.

Droplets were generated using the AutoDG Automated Droplet Generator (Bio-Rad, Hercules, CA). PCR was performed using Mastercycler Pro (Eppendforf, Enfield, CT) with the following cycling conditions: reverse transcription at 50 °C for 60 min, enzyme activation at 95 °C for 5 min, 40 cycles of denaturation at 95 °C for 30 s and annealing and extension at 59 °C (for HIV-1) or 56 °C (for PMMoV) for 30 s, enzyme deactivation at 98 °C for 10 min then an indefinite hold at 4 °C. The ramp rate for temperature changes were set to 2 °C/second and the final hold at 4 °C was performed for a minimum of 30 min to allow the droplets to stabilize. Droplets were analyzed using the QX200 or the QX600 Droplet Reader (Bio-Rad). A well had to have over 10,000 droplets for inclusion in the analysis. All liquid transfers were performed using the Agilent Bravo (Agilent Technologies, Santa Clara, CA).

Thresholding was done using QuantaSoft™ Analysis Pro Software (Bio-Rad, version 1.0.596) for data generated using the QX200 and QX Manager Software (Bio-Rad, version 2.0) for data generated using the QX600. In order for a sample to be recorded as positive, it had to have at least 3 positive droplets. Replicate wells were merged for analysis of each sample.

Dimensional analysis was used to convert concentrations of nucleic-acid targets from copies per reaction to copies per dry weight of solids (cp/g) or copies per milliliter (cp/ml) of influent. Measurement errors are reported as standard deviations. The lowest detectable concentrations were ∼500-1000 cp/g for solids and 1 cp/ml for liquids, respectively, and is equivalent to the concentration giving rise to three positive droplets across 10 merged well. The range provided for solids reflects differences in the dry weight of different solids samples. Wastewater data for the retrospective study are publicly available through the Stanford Digital Repository (https://doi.org/10.25740/yz257qj0009).

### Statistics

A Wilcoxon rank sum test was used to determine if there were significant differences in the concentrations of nucleic acids between the two utilities with the alternative hypothesis that concentrations were higher at OSP than SJ. A Wilcoxon signed rank test was used to determine whether there were significant differences in the concentration estimated between matched samples tested with and without a reverse transcription step. A Wilcoxon signed rank test was also used to determine whether there were significant differences in the concentration estimated on a per mass basis (cp/g and cp/mL) between matched liquid and solids samples.

### Clinical data

Estimated annual numbers of incident cases of HIV were obtained at the county level from publicly available county health department surveillance summaries for 2021 and 2022^5,35–37^. Estimates of PLWH and the proportions with viral suppression were also obtained from these reports. Data for 2023 was not available at the time of writing; further details of health department reporting and definitions are available in the SI. Rates of PLWH per county were calculated based on yearly population denominators obtained from the State of California, Department of Finance, Demographic Research Unit^38^.

## Results

### Quality control

Positive and negative extraction and PCR controls were positive and negative, respectively indicating assays performed and no evidence of contamination. The PMMoV endogenous control, as expected, was detected at relatively high concentrations in wastewater (median (range) log_10_-transformed PMMoV = 9.1 (8.5-10.2) and 8.7 (8.1-9.5) log_10_ copies/g at SJ and OSP, respectively). Given that the lowest measurements at the two sites were within an order of magnitude of the medians, we concluded that there was no gross extraction failure. Further details of the methods and controls per the Environmental Microbiology Minimal Information (EMMI) guidelines can be found in the supporting materials (Figure S2).

We compared PMMoV measured using the biobanked and stored samples to PMMoV measured in the same samples when processed immediately for a routine wastewater surveillance project As described in Boehm et al.^39^, median (interquartile range) ratio of PMMoV RNA measurements made in this study to those made using fresh samples was 0.9 (0.6-1.2) at SJ and 1.0 (0.7-1.4) at OSP suggesting limited degradation of the PMMoV RNA target in the stored and freeze thawed samples. It should be noted that there could have still been degradation of HIV-1 nucleic-acids in these biobanked samples, especially since the N gene from SARS-CoV-2 has shown some losses under these storage conditions^39^; however, sample storage is essential for retrospective work.

### Analysis of HIV-1 in wastewater

HIV-1 nucleic-acids (NA) were detected consistently throughout the retrospective study period in both OSP and SJ wastewater (Figure 1). HIV-1 NA were detected in 94% (215/230) of samples from OSP and 23% (53/229) of samples from SJ. Measured HIV-1 NA concentrations were significantly higher at OSP compared to SJ (Wilcoxon rank sum test, p<10^−16^, Figure 1). At OSP, concentrations ranged from non-detect to 3.9×10^5^ cp/g with a median 7.3×10^3^ cp/g. At SJ, concentrations ranged from non-detect to 1.1×10^5^ cp/g with a median of non-detect.

**Fig 1.**
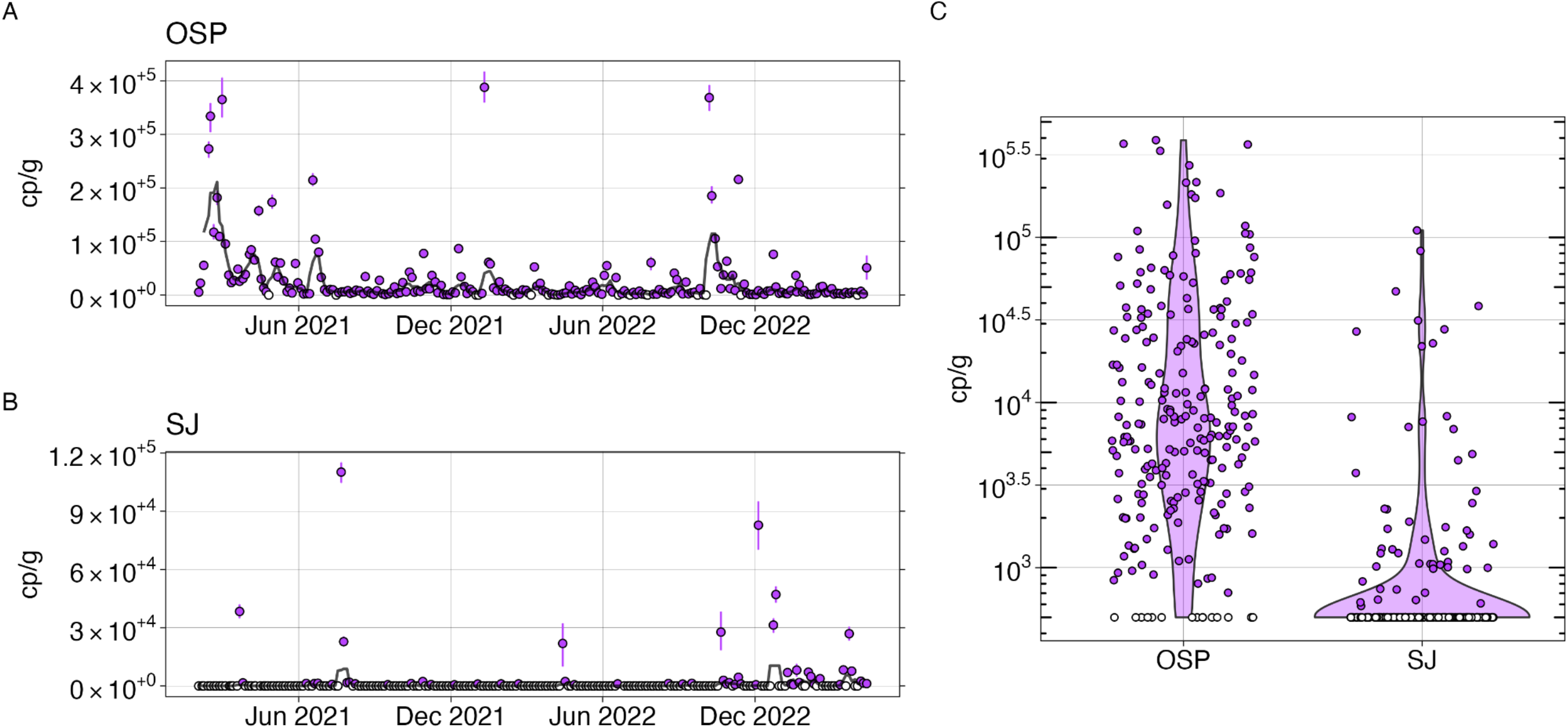
Wastewater concentrations of HIV nucleic-acids. **Left:** Concentration of HIV nucleic-acids in wastewater solids from OSP (A) and SJ (B) expressed in terms of copies of the target per gram dry weight of solids found in wastewater. Open circles show where HIV nucleic-acids were not detected, and the errors on each point represent standard deviations (68% confidence intervals) from the droplet digital RT-PCR measurement and include Poisson error as well as variation among replicate wells. The black line represents the 5-sample smoothed and trimmed average. **Right**: Distribution of the concentration of HIV nucleic-acids in wastewater solids from OSP and SJ (C), in terms of copies of the target per gram dry weight of solids found in wastewater. Open circles show where HIV nucleic-acids were not detected.

Measured HIV-1 NA concentrations were significantly different in solids compared to liquids (Wilcoxon signed rank test, p < 10^−3^). Concentrations in solids were enriched by several orders of magnitude compared to liquids on a per mass basis with a median concentration of 1 cp/mL in liquids and 6142 cp/g in solids across all samples included in the comparison (Fig 2). Liquid samples also had a higher rate of nondetection for HIV-1; HIV-1 NA were not detected in 4/16 liquids samples and 2/16 solids samples. One sample had no HIV-1 NA detected by either method.

**Fig 2.**
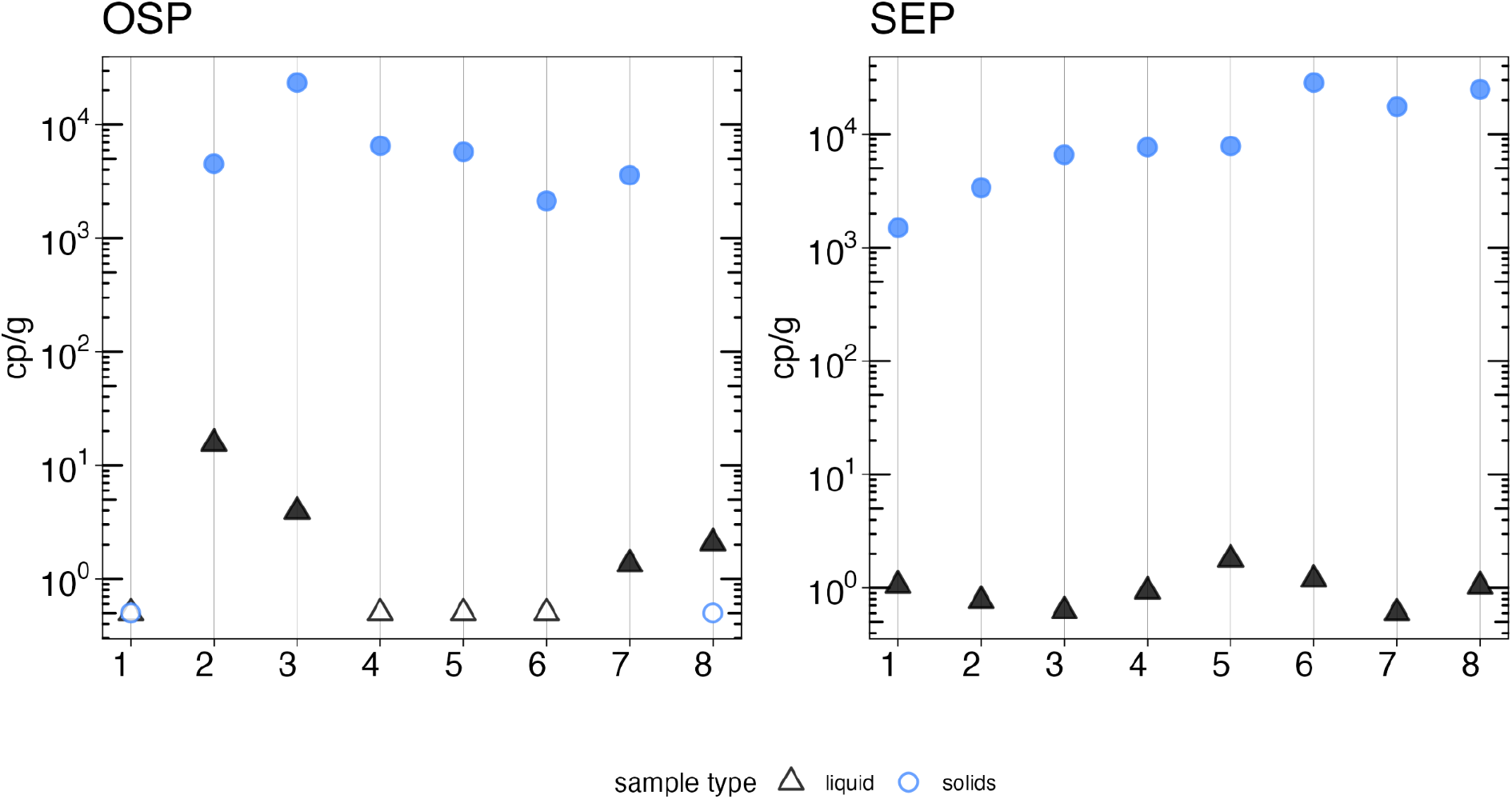
Comparison of HIV nucleic acid quantification in liquids and solids. Concentrations of HIV nucleic acids in wastewater liquids (black triangles) and solids (blue circles) from OSP and SEP expressed in terms of copies of the target per mL liquid wastewater or grams dry weight of solids found in wastewater. Open circles or triangles show where HIV nucleic acids were not detected.

When analyzed with and without a reverse transcription step, HIV-1 NA in wastewater were significantly different (Wilcoxon signed rank test, p = 0.03) but the magnitude of the difference between the two was small (Fig 3; with RT median = 5773 cp/g and mean = 8734 cp/g, without RT median = 8012 cp/g and mean = 10296 cp/g).

**Fig 3.**
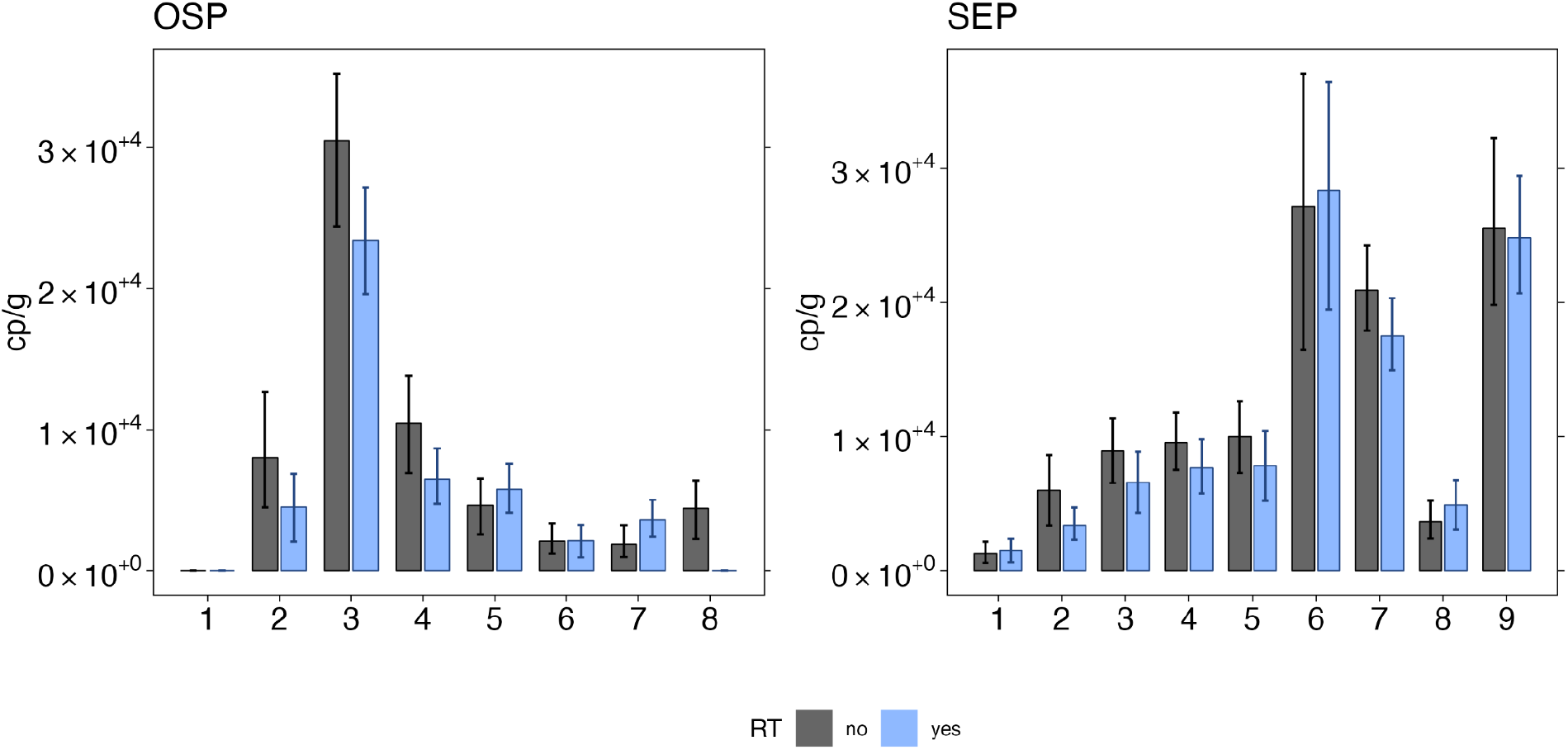
Comparison of HIV nucleic acid quantification with and without RT. Concentrations of HIV nucleic acids in wastewater solids from OSP and SEP are expressed in terms of copies of the target per gram dry weight of solids found in wastewater for quantification with the use of RT (blue) and without (black). Errors on each bar represent standard deviations (68% confidence intervals) from the droplet digital RT-PCR measurement and include Poisson error as well as variation among replicate wells.

### Clinical Data

The number of new diagnoses of HIV from San Francisco was similar to the number of new diagnoses in Santa Clara County for both 2021 and 2022 (Table 2), however as the population of Santa Clara County is much larger, the rate of new diagnoses per 100,000 residents was higher in SF than in SCC. The number and rate of PLWH was also higher in San Francisco County compared to Santa Clara County. The two counties have similar rates of viral suppression, estimated at 72-73% for San Francisco and 70.2-68.1% for Santa Clara County from 2021 - 2022.

**Table 2.** Numbers and rates of PLWH, %PLWH who were virally suppressed, and new HIV diagnoses in San Francisco and Santa Clara Counties in 2021 and 2022.

|  | San Francisco County |  | Santa Clara County |  |
| --- | --- | --- | --- | --- |
|  | 2021 | 2022 | 2021 | 2022 |
| **Total PLWH, N** | 11830 | 11798 | 3618 | 3770 |
| **PLWH per 100,000 population\*** | 1405.5 | 1399.4 | 190.4 | 198.1 |
| **New diagnoses, N** | 166 | 157 | 133 | 165 |
| **New diagnoses per 100 000 population\*** | 19.7 | 18.6 | 7.0 | 8.7 |
| **Viral suppression, %** | 72% | 73% | 70.2% | 68.1% |

These county level HIV prevalence estimates serve as imperfect proxies for the population living in the OSP and SJ sewersheds. OSP WWTP serves 25% of the population of San Francisco county, and annual reports show that the geographic area served by the plant has lower rates of PLWH than the county as a whole^37,40^. Neighborhoods within the OSP service area are described as having a rate of 317-999/100,000 PLWH in 2021, and 310-799/100,000 PLWH in 2022. SJ WWTP serves 75% of the population of Santa Clara County.

### Comparison of clinical and wastewater data

Overall, the rate of detection and concentrations of HIV-1 nucleic acids were higher in San Francisco County than in Santa Clara County and the rates of new HIV diagnoses and rates of PLWH were also higher in San Francisco County. No statistics are applied to this comparison as clinical data is available at an annual scale and N = 4, limiting appropriate hypothesis testing.

## Discussion

This study demonstrates a first proof of concept that HIV-1 nucleic-acids can be reliably detected and quantified in wastewater, and rates of detection and concentration are aligned with known information about the overall prevalence of PLWH in the associated communities. HIV-1 nucleic acids were consistently detected in wastewater from WWTPs in the San Francisco Bay Area, California (CA), USA, with differences in wastewater rates of detection concentrations that mirrored clinical surveillance data between counties. Both the rate of detection and concentrations of HIV-1 nucleic acids were higher in San Francisco County than in Santa Clara County, consistent with the higher rate of both PLWH and new HIV diagnoses in San Francisco. Although the sewershed area served by the San Francisco County WWTP covers a part of the city that has relatively lower prevalence of residents with PLWH, even these areas have reported prevalence that is approximately double or more the prevalence in Santa Clara County.

HIV-1 nucleic acids were detected from both the liquid and solid portion of wastewater from two sites in San Francisco County. The concentrations in solids were several orders of magnitude higher on a per mass basis than those in liquid, and the rate of non-detection was slightly higher in liquid samples. This suggests that HIV-1 nucleic acids can be successfully quantified in wastewater utilizing either the solid or the liquid matrix, but the use of solids may improve the sensitivity of the measurements. Future work utilizing liquids and solids across a wider range of anticipated concentration of HIV-1 NA would help determine the performance of methods utilizing each matrix at different levels.

The field of wastewater monitoring for infectious disease has grown rapidly in recent years and has not only become a part of many public health department’s approach to infectious disease surveillance, it has been successfully used to monitor diseases like mpox that interact with HIV. Wastewater monitoring has not, however, been previously used to support HIV monitoring.This is likely because HIV has a number of important differences from other viruses commonly monitored using wastewater such as respiratory and enteric viruses, particularly when considering the possibility of long term shedding and how this relates to individuals with different disease outcomes^19,20^.

There are limited quantitative data on the concentration of HIV-1 nucleic-acids in excretions. While individuals with active infections may shed viral RNA, PLWH who are virally suppressed may still excrete HIV-1 DNA^19^. In this study, we quantified total HIV-1 nucleic-acids which include both RNA and DNA; the methods we used cannot distinguish between the two. The overall similarity in concentrations of HIV nucleic-acids with and without reverse transcription in a small subset of samples suggests that the majority of the nucleic-acids detected in wastewater are DNA. Further work will need to be done to better understand shedding dynamics of HIV-1 nucleic-acids in PLWH and those with viremia to provide a foundation to further interpret these results.

This work presents a first proof of concept; in order to make HIV monitoring in wastewater a tool that is useful for public health response to the HIV epidemic more work is necessary to understand the relationship between these measurements and PLWH and even more importantly the people living with undiagnosed or unmanaged HIV in the community. Multiple assays in the future might be used to distinguish active HIV replication from the proviral reservoir, both of which may be represented by nucleic acids in wastewater, an increase in cases that are untreated vs effectively treated may be detectable. Targets in wastewater that identify HIV resistance may help inform initial antiretroviral therapy (ART) choices (e.g., is the virus mutating, is pre-exposure prophylaxis (PrEP) contributing to resistance, etc.) without identifying or linking individuals.

Despite CDC recommendations for routine HIV screening in healthcare settings since 2006^41^, HIV testing rates during ambulatory care visits have remained low^7,42^. With barriers to implementation ranging from provider perceptions of low HIV risk to institutional concerns surrounding allocation of limited resources^7,42^, recent initiatives, such as the EHE, propose refocused efforts to target areas with the highest burden of HIV ^43,44^. With more information from studies on HIV NA shedding and a closer comparison of HIV infections and wastewater data to guide interpretation of presence and concentrations of HIV NA targets in wastewater and the relationship to infection and disease outcomes, data on HIV from wastewater may be useful both at a facility and community level for monitoring trends to help inform screening efforts.

Tools like this would provide opportunities to compare the burden of HIV across geographic areas, to provide information on the scope of the HIV burden in institutions like hospitals and correctional facilities, and possibly to determine patterns in resistance. This may be used to determine times and places when increased screening at sites may be useful, feeding into existing frameworks that identify priority areas for intervention.

## Data Availability

All data produced are available online at the Stanford Digital Repository.

https://doi.org/10.25740/yz257qj0009

## Acknowledgements

We acknowledge Gregory Ostolaza Bravo for contributions to wastewater sample analysis.

## Notes

### Competing Interest Statement

Dorothea Duong, Bridgette Shelden and Bradley White are employees of Verily Life Sciences, LLC

### Funding Statement

This study was funded in part by National Institute of Health (R21-AI179550) and the Sergey Brin Family Foundation.

### Author Declarations

The study used ONLY openly available human data that were originally located at https://www.cdc.gov/hiv/library/reports/index.html, https://publichealth.sccgov.org/sites/g/files/exjcpb916/files/documents/STIHIV_AnnualReport_2022.pdf, https://publichealth.sccgov.org/sites/g/files/exjcpb916/files/documents/STIHIV_AnnualReport_2021.pdf, https://www.sfdph.org/dph/files/reports/RptsHIVAIDS/AnnualReport2022-Orange.pdf

## References

(1) Why the HIV epidemic is not over. https://www.who.int/news-room/spotlight/why-the-hiv-epidemic-is-not-over (accessed 2024-01-29).

(2) HIV Surveillance | Reports| Resource Library | HIV/AIDS | CDC. https://www.cdc.gov/hiv/library/reports/hiv-surveillance.html (accessed 2024-03-08).

(3) HIV Surveillance | Reports| Resource Library | HIV/AIDS | CDC. https://www.cdc.gov/hiv/library/reports/hiv-surveillance.html (accessed 2024-03-08).

(4) CDC-Funded HIV Testing in the United States, Puerto Rico, and the U.S. Virgin Islands, 2020.; Centers for Disease Control and Prevention, 2022. https://www.cdc.gov/hiv/library/reports/index.html (accessed 2024-03-10).

(5) CDC-Funded HIV Testing in the United States, Puerto Rico, and the U.S. Virgin Islands, 2021.; Centers for Disease Control and Prevention, 2023. https://www.cdc.gov/hiv/library/reports/index.html (accessed 2024-03-10).

(6) Estimated HIV Incidence and Prevalence in the United States, 2017–2021. HIV Surveillance Supplemental Report, 2023; 28 (No.3); Centers for Disease Control and Prevention, 2023. https://www.cdc.gov/hiv/library/reports/index.html (accessed 2024-03-10).

(7) Hoover, K. W. HIV Services and Outcomes During the COVID-19 Pandemic — United States, 2019–2021. MMWR Morb. Mortal. Wkly. Rep. 2022, 71. 10.15585/mmwr.mm7148a1.

(8) Boehm, A. B.; Hughes, B.; Duong, D.; Chan-Herur, V.; Buchman, A.; Wolfe, M. K.; White, B. J. Wastewater Concentrations of Human Influenza, Metapneumovirus, Parainfluenza, Respiratory Syncytial Virus, Rhinovirus, and Seasonal Coronavirus Nucleic-Acids during the COVID-19 Pandemic: A Surveillance Study. Lancet Microbe 2023, 4 (5). 10.1016/S2666-5247(22)00386-X.

(9) Peccia, J.; Zulli, A.; Brackney, D. E.; Grubaugh, N. D.; Kaplan, E. H.; Casanovas-Massana, A.; Ko, A. I.; Malik, A. A.; Wang, D.; Wang, M.; Warren, J. L.; Weinberger, D. M.; Omer, S. B. SARS-CoV-2 RNA Concentrations in Primary Municipal Sewage Sludge as a Leading Indicator of COVID-19 Outbreak Dynamics; preprint; Epidemiology, 2020. 10.1101/2020.05.19.20105999.

(10) Wolfe, M. K.; Yu, A. T.; Duong, D.; Rane, M. S.; Hughes, B.; Chan-Herur, V.; Donnelly, M.; Chai, S.; White, B. J.; Vugia, D. J.; Boehm, A. B. Use of Wastewater for Mpox Outbreak Surveillance in California. N. Engl. J. Med. 2023, 388 (6), 570–572. 10.1056/NEJMc2213882.

(11) Boehm, A. B.; Wolfe, M. K.; White, B. J.; Hughes, B.; Duong, D.; Banaei, N.; Bidwell, A. Human Norovirus (HuNoV) GII RNA in Wastewater Solids at 145 United States Wastewater Treatment Plants: Comparison to Positivity Rates of Clinical Specimens and Modeled Estimates of HuNoV GII Shedders. J. Expo. Sci. Environ. Epidemiol. 2023, 1–8. 10.1038/s41370-023-00592-4.

(12) Boehm, A. B.; Hughes, B.; Duong, D.; Banaei, N.; White, B. J.; Wolfe, M. K. A Retrospective Longitudinal Study of Adenovirus Group F, Norovirus GI and GII, Rotavirus, and Enterovirus Nucleic-Acids in Wastewater Solids at Two Wastewater Treatment Plants: Solid-Liquid Partitioning and Relation to Clinical Testing Data. medRxiv August 31, 2023, p 2023.08.29.23294748. 10.1101/2023.08.29.23294748.

(13) Wolfe, M. K.; Topol, A.; Knudson, A.; Simpson, A.; White, B.; Vugia, D. J.; Yu, A. T.; Li, L.; Balliet, M.; Stoddard, P.; Han, G. S.; Wigginton, K. R.; Boehm, A. B. High-Frequency, High-Throughput Quantification of SARS-CoV-2 RNA in Wastewater Settled Solids at Eight Publicly Owned Treatment Works in Northern California Shows Strong Association with COVID-19 Incidence. mSystems 6 (5), e00829–21. 10.1128/mSystems.00829-21.

(14) Wolfe, M. K.; Duong, D.; Bakker, K. M.; Ammerman, M.; Mortenson, L.; Hughes, B.; Arts, P.; Lauring, A. S.; Fitzsimmons, W. J.; Bendall, E.; Hwang, C. E.; Martin, E. T.; White, B. J.; Boehm, A. B.; Wigginton, K. R. Wastewater-Based Detection of Two Influenza Outbreaks. Environ. Sci. Technol. Lett. 2022, 9 (8), 687–692. 10.1021/acs.estlett.2c00350.

(15) Preston, D. R.; Farrah, S. R.; Bitton, G.; Chaudhry, G. R. Detection of Nucleic Acids Homologous to Human Immunodeficiency Virus in Wastewater. J. Virol. Methods 1991, 33 (3), 383–390. 10.1016/0166-0934(91)90038-2.

(16) Ansari, S. A.; Farrah, S. R.; Chaudhry, G. R. Presence of Human Immunodeficiency Virus Nucleic Acids in Wastewater and Their Detection by Polymerase Chain Reaction. Appl. Environ. Microbiol. 1992, 58 (12), 3984–3990.

(17) Casson, L. W.; Ritter, M. O. D.; Cossentino, L. M.; Gupta, P. Survival and Recovery of Seeded HIV in Water and Wastewater. Water Environ. Res. 1997, 69 (2), 174–179.

(18) Moore, B. E. Survival of Human Immunodeficiency Virus (HIV), HIV-Infected Lymphocytes, and Poliovirus in Water. Appl. Environ. Microbiol. 1993, 59 (5), 1437–1443. 10.1128/aem.59.5.1437-1443.1993.

(19) Chakrabarti, A. K.; Caruso, L.; Ding, M.; Shen, C.; Buchanan, W.; Gupta, P.; Rinaldo, C. R.; Chen, Y. Detection of HIV-1 RNA/DNA and CD4 mRNA in Feces and Urine from Chronic HIV-1 Infected Subjects with and without Anti-Retroviral Therapy. AIDS Res. Ther. 2009, 6, 20. 10.1186/1742-6405-6-20.

(20) van der Hoek, L.; Boom, R.; Goudsmit, J.; Snijders, F.; Sol, C. J. Isolation of Human Immunodeficiency Virus Type 1 (HIV-1) RNA from Feces by a Simple Method and Difference between HIV-1 Subpopulations in Feces and Serum. J. Clin. Microbiol. 1995, 33 (3), 581–588. 10.1128/jcm.33.3.581-588.1995.

(21) Zuckerman, R. A.; Whittington, W. L. H.; Celum, C. L.; Collis, T. K.; Lucchetti, A. J.; Sanchez, J. L.; Hughes, J. P.; Sanchez, J. L.; Coombs, R. W. Higher Concentration of HIV RNA in Rectal Mucosa Secretions than in Blood and Seminal Plasma, among Men Who Have Sex with Men, Independent of Antiretroviral Therapy. J. Infect. Dis. 2004, 190 (1), 156–161. 10.1086/421246.

(22) Wolfe, M. K.; Topol, A.; Knudson, A.; Simpson, A.; White, B.; Duc, V.; Yu, A.; Li, L.; Balliet, M.; Stoddard, P.; Han, G.; Wigginton, K. R.; Boehm, A. High-Frequency, High-Throughput Quantification of SARS-CoV-2 RNA in Wastewater Settled Solids at Eight Publicly Owned Treatment Works in Northern California Shows Strong Association with COVID-19 Incidence. mSystems 2021, 6 (5), e00829-21. 10.1128/mSystems.00829-21.

(23) Topol, A.; Wolfe, M.; White, B.; Wigginton, K.; Boehm, A. High Throughput Pre-Analytical Processing of Wastewater Settled Solids for SARS-CoV-2 RNA Analyses. protocols.io 2021. 10.17504/protocols.io.btyqnpvw.

(24) Boehm, A. B.; Wolfe, M. K.; Wigginton, K. R.; Bidwell, A.; White, B. J.; Hughes, B.; Duong; Chan-Herur, V.; Bischel, H. N.; Naughton, C. C. Human Viral Nucleic Acids Concentrations in Wastewater Solids from Central and Coastal California USA. Sci. Data 2023, 10, 396.

(25) Topol, A.; Wolfe, M.; Wigginton, K.; White, B.; Boehm, A. High Throughput RNA Extraction and PCR Inhibitor Removal of Settled Solids for Wastewater Surveillance of SARS-CoV-2 RNA. protocols.io 2021. 10.17504/protocols.io.btyrnpv6.

(26) Visseaux, B.; Damond, F.; Matheron, S.; Descamps, D.; Charpentier, C. Hiv-2 Molecular Epidemiology. Infect. Genet. Evol. 2016, 46, 233–240. 10.1016/j.meegid.2016.08.010.

(27) Kibirige, C. N.; Manak, M.; King, D.; Abel, B.; Hack, H.; Wooding, D.; Liu, Y.; Fernandez, N.; Dalel, J.; Kaye, S.; Imami, N.; Jagodzinski, L.; Gilmour, J. Development of a Sensitive, Quantitative Assay with Broad Subtype Specificity for Detection of Total HIV-1 Nucleic Acids in Plasma and PBMC. Sci. Rep. 2022, 12 (1), 1550. 10.1038/s41598-021-03016-1.

(28) Brussel, A.; Delelis, O.; Sonigo, P. Alu-LTR Real-Time Nested PCR Assay for Quantifying Integrated HIV-1 DNA. In Human Retrovirus Protocols: Virology and Molecular Biology; Zhu, T., Ed.; Methods in Molecular BiologyTM; Humana Press: Totowa, NJ, 2005; pp 139–154. 10.1385/1-59259-907-9:139.

(29) Friedrich, B.; Li, G.; Dziuba, N.; Ferguson, M. R. Quantitative PCR Used to Assess HIV-1 Integration and 2-LTR Circle Formation in Human Macrophages, Peripheral Blood Lymphocytes and a CD4+ Cell Line. Virol. J. 2010, 7 (1), 354. 10.1186/1743-422X-7-354.

(30) McClary-Gutierrez, J. S.; Aanderud, Z. T.; Al-faliti, M.; Duvallet, C.; Gonzalez, R.; Guzman, J.; Holm, R. H.; Jahne, M. A.; Kantor, R. S.; Katsivelis, P.; Kuhn, K. G.; Langan, L. M.; Mansfeldt, C.; McLellan, S. L.; Mendoza Grijalva, L. M.; Murnane, K. S.; Naughton, C. C.; Packman, A. I.; Paraskevopoulos, S.; Radniecki, T. S.; Roman, F. A.; Shrestha, A.; Stadler, L. B.; Steele, J. A.; Swalla, B. M.; Vikesland, P.; Wartell, B.; Wilusz, C. J.; Wong, J. C. C.; Boehm, A. B.; Halden, R. U.; Bibby, K.; Delgado Vela, J. Standardizing Data Reporting in the Research Community to Enhance the Utility of Open Data for SARS-CoV-2 Wastewater Surveillance. Environ. Sci. Water Res. Technol. 2021, 7 (9), 1545–1551. 10.1039/D1EW00235J.

(31) Kantor, R. S.; Nelson, K. L.; Greenwald, H. D.; Kennedy, L. C. Challenges in Measuring the Recovery of SARS-CoV-2 from Wastewater. Environ. Sci. Technol. 2021, 55 (6), 3514–3519. 10.1021/acs.est.0c08210.

(32) Boehm, A. B.; Wolfe, M. K.; White, B. J.; Hughes, B.; Duong, D.; Bidwell, A. More than a Tripledemic: Influenza A Virus, Respiratory Syncytial Virus, SARS-CoV-2, and Human Metapneumovirus in Wastewater during Winter 2022–2023. Environ. Sci. Technol. Lett. 2023, 10 (8), 622–627. 10.1021/acs.estlett.3c00385.

(33) Boehm, A. B.; Wolfe, M. K.; White, B. J.; Hughes, B.; Duong, D. Two Years of Longitudinal Measurements of Human Adenovirus Group F, Norovirus GI and GII, Rotavirus, Enterovirus, Enterovirus D68, Hepatitis A Virus, Candida Auris, and West Nile Virus Nucleic-Acids in Wastewater Solids: A Retrospective Study at Two Wast…. medRxiv 2023. 10.1101/2023.08.22.23294424.

(34) Topol, A.; Wolfe, M.; White, B.; Wigginton, K.; Boehm, A. High Throughput SARS-COV-2, PMMOV, and BCoV Quantification in Settled Solids Using Digital RT-PCR. protocols.io 2021. 10.17504/protocols.io.btywnpxe.

(35) Sexually Transmitted Infections (STI) and HIV Epidemiology Annual Report, 2022; County of Santa Clara Public Health Department Infectious Disease and Response Branch, 2023. https://publichealth.sccgov.org/sites/g/files/exjcpb916/files/documents/STIHIV_AnnualReport_2022.pdf (accessed 2024-03-10).

(36) Sexually Transmitted Infections (STI) and HIV Epidemiology Annual Report, 2021; County of Santa Clara Public Health Department Infectious Disease and Response Branch, 2023. https://publichealth.sccgov.org/sites/g/files/exjcpb916/files/documents/STIHIV_AnnualReport_2021.pdf (accessed 2024-03-10).

(37) HIV Epidemiology Annual Report 2022; San Francisco Department of Public Health Population Health Division HIV Epidemiology Section, 2023. https://www.sfdph.org/dph/files/reports/RptsHIVAIDS/AnnualReport2022-Orange.pdf(accessed 2024-03-10).

(38) E-2. California County Population Estimates and Components of Change by Year — July 1, 2020-2023 | Department of Finance; 2023. https://dof.ca.gov/forecasting/demographics/estimates/e-2/ (accessed 2024-03-11).

(39) Boehm, A. B.; Hughes, B.; Duong, D.; Banaei, N.; White, B. J.; Wolfe, M. K. A Retrospective Longitudinal Study of Adenovirus Group F, Norovirus GI and GII, Rotavirus, and Enterovirus Nucleic-Acids in Wastewater Solids at Two Wastewater Treatment Plants: Solid-Liquid Partitioning and Relation to Clinical Testing Data. Revis. MSphere 2024.

(40) HIV Epidemiology Annual Report 2021; San Francisco Department of Public Health Population Health Division HIV Epidemiology Section, 2022. https://www.sfdph.org/dph/files/reports/RptsHIVAIDS/AnnualReport2021-Red.pdf(accessed 2024-03-10).

(41) Branson, B. M.; Handsfield, H. H.; Lampe, M. A.; Janssen, R. S.; Taylor, A. W.; Lyss, S. B.; Clark, J. E.; Centers for Disease Control and Prevention (CDC). Revised Recommendations for HIV Testing of Adults, Adolescents, and Pregnant Women in Health-Care Settings. MMWR Recomm. Rep. Morb. Mortal. Wkly. Rep. Recomm. Rep. 2006, 55 (RR-14), 1–17; quiz CE1-4.

(42) Olatosi, B.; Siddiqi, K. A.; Conserve, D. F. Towards Ending the Human Immunodeficiency Virus Epidemic in the US. Medicine (Baltimore) 2020, 99 (2), e18525. 10.1097/MD.0000000000018525.

(43) Berg, L. J.; Delgado, M. K.; Ginde, A. A.; Montoy, J. C.; Bendavid, E.; Camargo, C. A. Characteristics of U.S. Emergency Departments That Offer Routine Human Immunodeficiency Virus Screening. Acad. Emerg. Med. Off. J. Soc. Acad. Emerg. Med. 2012, 19 (8), 894–900. 10.1111/j.1553-2712.2012.01401.x.

(44) Shirreffs, A.; Lee, D. P.; Henry, J.; Golden, M. R.; Stekler, J. D. Understanding Barriers to Routine HIV Screening: Knowledge, Attitudes, and Practices of Healthcare Providers in King County, Washington. PLOS ONE 2012, 7 (9), e44417. 10.1371/journal.pone.0044417.

