## Supporting Material for "Detection and quantification of human immunodeficiency virus-1 (HIV-1) total nucleic acids in wastewater settled solids from two California communities"

**Additional methods.** In California, healthcare providers and laboratories are required to report a confirmed HIV test to their local health jurisdiction. Specifically, data for Santa Clara County was obtained from the Santa Clara County Public Health "Sexually Transmitted Infections (STI) and HIV Epidemiology Annual Report"[1,2]. New HIV diagnoses are reported to Santa Clara County Public Health primarily from outpatient clinical settings, screening, diagnostic, and referral agencies, and inpatient clinical settings. The number of PLWH in the county is estimated based on people known to be living with HIV with a last known address in SCC, and the percentage of PLWH who are virally suppressed is based on the number of PLWH with a most recent viral load test less than 200 copies/ml in a given year. For San Francisco County, data was obtained from the San Francisco "HIV Epidemiology Annual Report" from the Department of Public Health, Population Health Division[3,4]. New HIV diagnoses are reported to San Francisco Public Health primarily through active surveillance of laboratory tests, pathology results and medical records. The number of PLWH in the county is estimated based on people know to be living with HIV with a last known address in San Francisco, and the percentage of PLWH who are virally suppressed is based on the number of PLWH with a most recent viral load test less than 200 copies/ml in the past 12 months.

**Additional details related to the EMMI guidelines.** Thirty-six samples from the retrospective study were selected at random for this analysis; this represents 8% of the samples processed in the study. As described in the methods section, each sample was run as template in two different PCR reactions; 1 for PMMoV, 1 for HIV-1, SARS-CoV-2, Influenza H1 gene, Influenza A N1 gene, West Nile Virus, Norovirus GII, human adenovirus group F, and rotavirus. The average (standard deviation) number of partitions (droplets) for each of the two reactions (across the 10 replicates) was 164164 (38851) for the reaction for PMMoV, 174321 (20126) for the reaction for HIV-1. The volume of the partitions, as reported by the machine vendor is 0.00085 µL. The mean and standard deviation of copies per partition for each target is shown in Table S3. Example fluorescent plots from the QX200 (two color reader) can be viewed in Topol et al. on protocols.io[5] and an example fluorescent plot from the QX600 (6 color reader) is included in the Stanford Digital Repository with the deposited data (https://doi.org/10.25740/yz257qj0009).

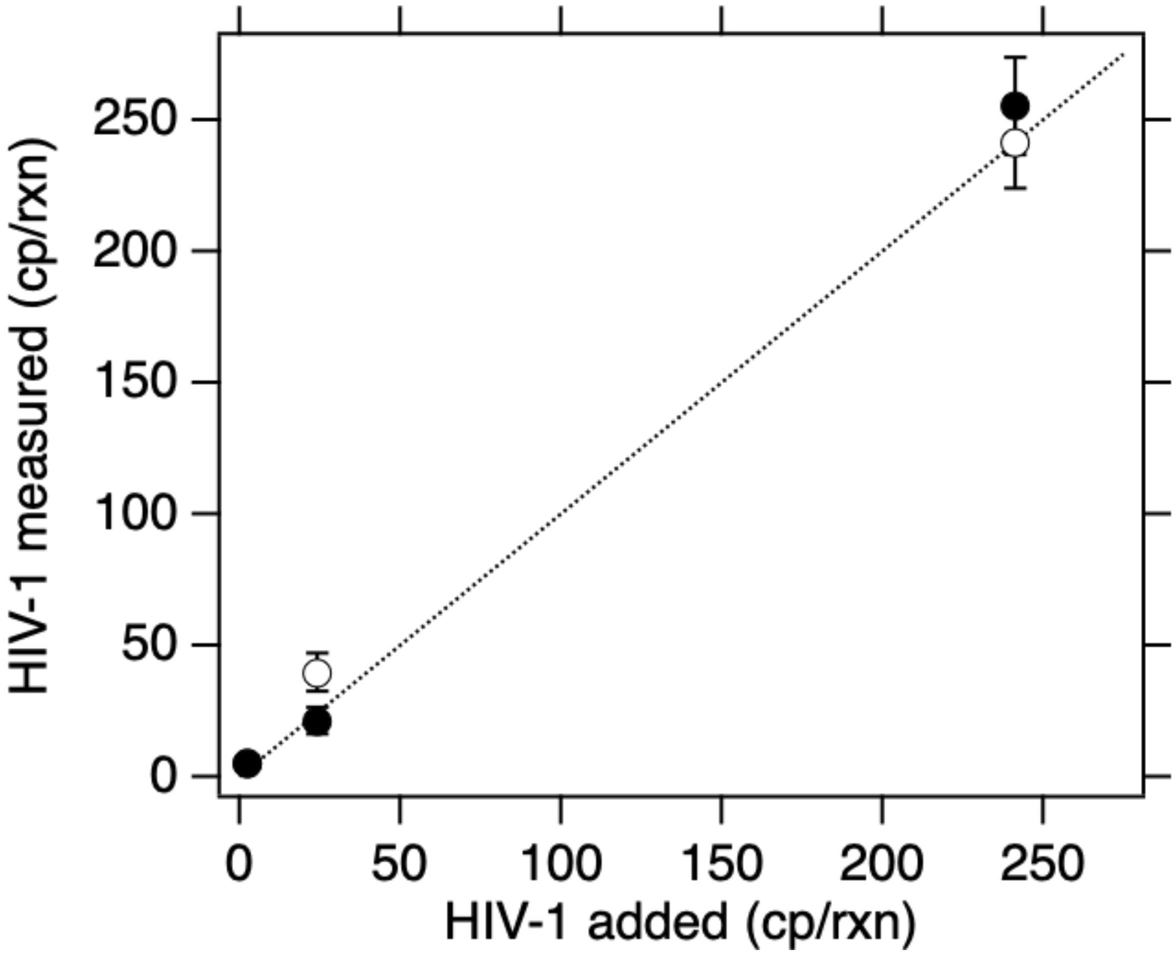


**Figure S1. Multiplex assay performance.** The concentration of the HIV-1 target in units of copies per reaction input and measured is provided. The white symbols are for reactions without the 7 background nucleic-acid targets and black symbols include high concentrations of the 7 other nucleic-acid targets. Error bars are standard deviations, if error bars cannot be seen, then they are smaller than the symbol. The line represents the 1:1 line.

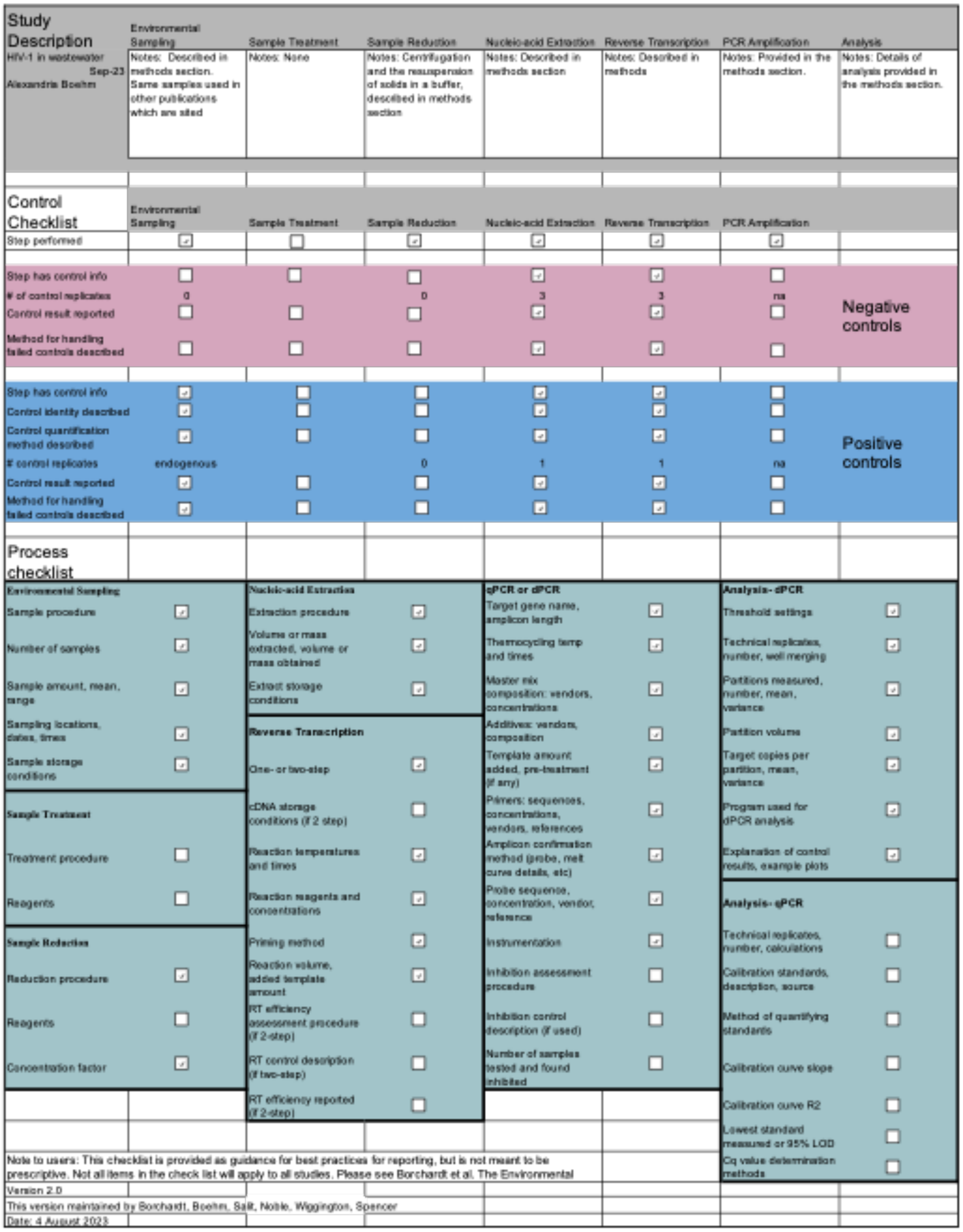


**Figure S2. EMMI[6] checklist.**

**Table S1. Sample collection details.**

| Wastewater plant | Dates of contemporaneous collection of liquids and solids | Dates of solids collection for analysis with and without RT step |
|---|---|---|
| **OSP** | July 11, 13-16, 17, 18, and 21, 2023 (n=8) | July 11, 13-16, 18, 19, 21, 2023 (n=9) |
| **SEP** | July 10-16, and 18, 2023 (n=8) | July 10-18, 2023 (n=9) |

**Table S2. Primers and probes for PMMoV.** Probes indicate which fluorescent molecular was used as well as the quencher. HEX, hexachloro-fluorescein;; ZEN, a proprietary internal quencher from Integrated DNA Technologies (Coralville, IA, USA); and IBFQ, Iowa Black FQ.

| Target | Primer/Probe | Sequence |
|---|---|---|
| PMMoV | Forward | GAGTGGTTTGACCTTAACGTTTGA |
| | Reverse | TTGTCGGTTGCAATGCAAGT |
| | Probe | CCTACCGAAGCAAATG (5' HEX/ZEN/3' IBFQ) |

**Table S3. Additional details related to the EMMI guidelines.** For each target measured in this study, the mean and standard deviation (sd) of the total number of copies of target per partition. Num is the number of samples out of a random 36 included in this analysis that had detectable target in them and thus contributed to the calculated mean and standard deviation.

| Target | HIV-1 | PMMoV |
|---|---|---|
| mean | 0.0002 | 0.15 |
| sd | 0.0003 | 0.069 |
| num | 18 | 36 |

### References


(1) *Sexually Transmitted Infections (STI) and HIV Epidemiology Annual Report, 2022*; County of Santa Clara Public Health Department Infectious Disease and Response Branch, 2023. https://publichealth.sccgov.org/sites/g/files/exjcpb916/files/documents/STIHIV_AnnualReport_2022.pdf (accessed 2024-03-10).

(2) *Sexually Transmitted Infections (STI) and HIV Epidemiology Annual Report, 2021*; County of Santa Clara Public Health Department Infectious Disease and Response Branch, 2023. https://publichealth.sccgov.org/sites/g/files/exjcpb916/files/documents/STIHIV_AnnualReport_2021.pdf (accessed 2024-03-10).

(3) *HIV Epidemiology Annual Report 2021*; San Francisco Department of Public Health Population Health Division HIV Epidemiology Section, 2022. https://www.sfdph.org/dph/files/reports/RptsHIVAIDS/AnnualReport2021-Red.pdf (accessed 2024-03-10).

(4) *HIV Epidemiology Annual Report 2022*; San Francisco Department of Public Health Population Health Division HIV Epidemiology Section, 2023. https://www.sfdph.org/dph/files/reports/RptsHIVAIDS/AnnualReport2022-Orange.pdf (accessed 2024-03-10).

(5) Topol, A.; Wolfe, M.; White, B.; Wigginton, K.; Boehm, A. High Throughput SARS-COV-2, PMMOV, and BCoV Quantification in Settled Solids Using Digital RT-PCR. *protocols.io* **2021**. https://doi.org/dx.doi.org/10.17504/protocols.io.btywnpxe.

(6) Borchardt, M. A.; Boehm, A. B.; Salit, M.; Spencer, S. K.; Wigginton, K. R.; Noble, R. T. The Environmental Microbiology Minimum Information (EMMI) Guidelines: qPCR and dPCR Quality and Reporting for Environmental Microbiology. *Environ. Sci. Technol.* **2021**, *55* (15), 10210–10223. https://doi.org/10.1021/acs.est.1c01767.
